# Modified Long-Axis In-Plane Ultrasound Technique Versus Conventional Palpation Technique For Radial Arterial Cannulation: A Prospective Randomized Controlled Trial

**DOI:** 10.1101/19001586

**Authors:** Jiebo Wang, Zhongmeng Lai, Xianfeng Weng, Yong Lin, Guohua Wu, Jiansheng Su, Qijian Huang, Jian Zeng, Junle Liu, Zisong Zhao, Ting Yan, Liangcheng Zhang, Linying Zhou

## Abstract

**Background:** A low first-pass success rate of radial artery cannulation was obtained when using the conventional palpation technique(C-PT) or ultrasound-guided techniques, we therefore evaluate the effect of a modified long-axis in-plane ultrasound technique (M-LAINUT) in guiding radial artery cannulation in adults.

**Methods:** We conducted a prospective, randomized and controlled clinical trial of 288 patients undergoing radial artery cannulation. Patients were randomized 1:1 to M-LAINUT or C-PT group at fujian medical university union hospital between 2017 and 2018. Radial artery cannulation was performed by three anesthesiologists with different experience. The outcome was the first and total radial artery cannulation success rates, the number of attempts and the cannulation time.

**Results:** 285 patients were statistically analyzed. The success rate of first attempt was 91.6% in the M-LAINUT group (n=143) and 57.7% in the C-PT group (n=142; P<0.001) (odds ratio, 7.9; 95% confidence interval, 4.0-15.7). The total success rate (≤5 min and ≤3 attempts) in the M-LAINUT group was 97.9%, compared to 84.5% in the palpation group (p <0.001) (odds ratio, 8.5; 95% confidence interval, 2.5-29.2). The total cannulation time was shorter and the number of attempts was fewer in the M-LAINUT group than that in the C-PT group (p <0. 05).

**Conclusion:** Modified long-axis in-plane ultrasound-guided radial artery cannulation can increase the first and total radial artery cannulation success rates, reduce the number of attempts and shorten the total cannulation time in adults.

## 1. Introduction

In the emergency room, intensive care unit (ICU) and operating room, especially in major operation and critical patients, arterial blood pressure monitoring is often carried out by arterial cannulation to monitor hemodynamic changes closely.

Radial artery cannulation was traditionally performed by palpation of radial artery pulsation. Ultrasound-guided techniques have been widely used in various arterial cannulations because of its visual characteristics and higher success rate of cannulation compared with traditional palpation.^1-3^

In recent years, an increasing number of studies have shown that the first success rate of ultrasound-guided radial artery cannulation is greater than traditional palpation and the success rate of cannulation is 68% to 100%, which mainly depends on the technique used, the skill level of the operator, the condition of the blood vessel, and the definition of successful puncture.^4-6^ Short-axis out-of-plane ultrasound guidance may have shortcomings, such as blurred local imaging, unclear pinpoint positioning and a high vascular penetration rate,^7^which make the first attempt cannulation success rate not very high.^2^ In the long-axis in-plane ultrasound guidance technique, two studies reported ultrasound success rates of 53% and 62%, respectively.^4,8^

Therefore, we designed a prospective, randomized and controlled clinical research method to evaluate the effect of the modified long-axis in-plane ultrasound technique(M-LAINUT) in guiding radial artery cannulation. We hypothesized the M-LAINUT would have a higher rate of cannulation on the attempt compared with the conventional palpation technique(C-PT).

## 2. Methods

The trial was registered prior to patient enrollment at clinicaltrials.gov (http://www.chictr.org.cn/index.aspx, number:ChiCTR-IOR-17011474;Principal investigator: Jiebo Wang, Date of registration: 23 May 2017).

Adult patients with the diameter of the radial artery was not less than 2.2 mm scheduled for elective surgery were included in this trial. Exclusion criteria were ulnar artery occlusion, history of forearm surgery, skin infection at the puncture site, abnormal modified Allen test, and coagulation dysfunction Randomization and Blinding

Patients were randomized 1:1 to M-LAINUT or the C-PT for the success rate of the attempt at radial artery cannulation. Before the study began, all the random numbers were generated by a computer and placed in sealed envelopes. Study assignment was concealed until after the decision had been made to radial artery cannulation and the patient was enrolled in the trial.

Three anesthesiologists who performed radial artery cannulation were divided into three categories according to the years of clinical training in anesthesia after graduation from medical college: one year (CA1), three years (CA3), and five years (CA5). At least 15 cases of radial artery catheterization guided by the M-LAINUT were performed for each operator before starting the study.

Before the patient was sent to the operating room, the diameter and depth of the radial artery were measured by a specialist anesthesiologist who did not perform artery cannulation using an ultrasound instrument (SonoSite M-Turbo Color Doppler Ultrasound Diagnostic instrument, Linear Array probe (L25N/13–6; SonoSite Inc)). All radial artery cannulations was performed before or after induction of general anesthesia according to anesthesiologist’s preference. The specified technique was then used to place the radial artery catheter using a 20-gauge intravenous catheter (Insyte; Becton Dickinson, LTD. USA). Towels and ultrasonic probe covers were provided if modified ultrasound was selected.

In the M-LAINUT group, the wrist joint angle of the patient was adjusted to 45°. The operator faced the end of the patient’s arm, and the probe was held in the left hand with the bracket as the supporting point, then the needle was held with right thumb and index finger just like pen-holding. After examining the artery in cross section, the US probe was rotated 90° to obtain the view of long axis artery (Figure 1A). The ultrasonic plane should be aligned with the long axis of the artery. The imaging mode of the ultrasound was switched to color doppler mode (CFM) to obtain the arterial blood flow signal. As the probe was slided slowly parallel to the long axis of artery, more abundant signal of the arterial blood flow could be obtained. (Figure 1B). The needle tip was placed on the contact point of the centre line of probe and the skin, and then inserted to the skin at an angle of 15°–40°. The needle should be always aligned with the centre line of the probe during the procedures of cannulation (Figure 1A). Once the needle tip compressed the anterior wall of the radial artery, the collapsed radial artery and the decrease of blood flow signal could be shown on the ultrasound image (Figure 1B). After confirming the tip of needle within the lumen of artery on the ultrasound image and the appearance of the arterial blood in the hub of needle, keeping continuous clear visualization of needle tip and the artery, the needle was advanced approximately 3-5mm. If the blood continued to flow into the hub, the needle was withdrawn 1 -2cm out of cannula, then the needle and cannula were pushed into the blood vessel, at last, the needle was pulled out (Figure 1C) and the cannula was connected to the arterial invasive monitoring system.

**Figure 1.**
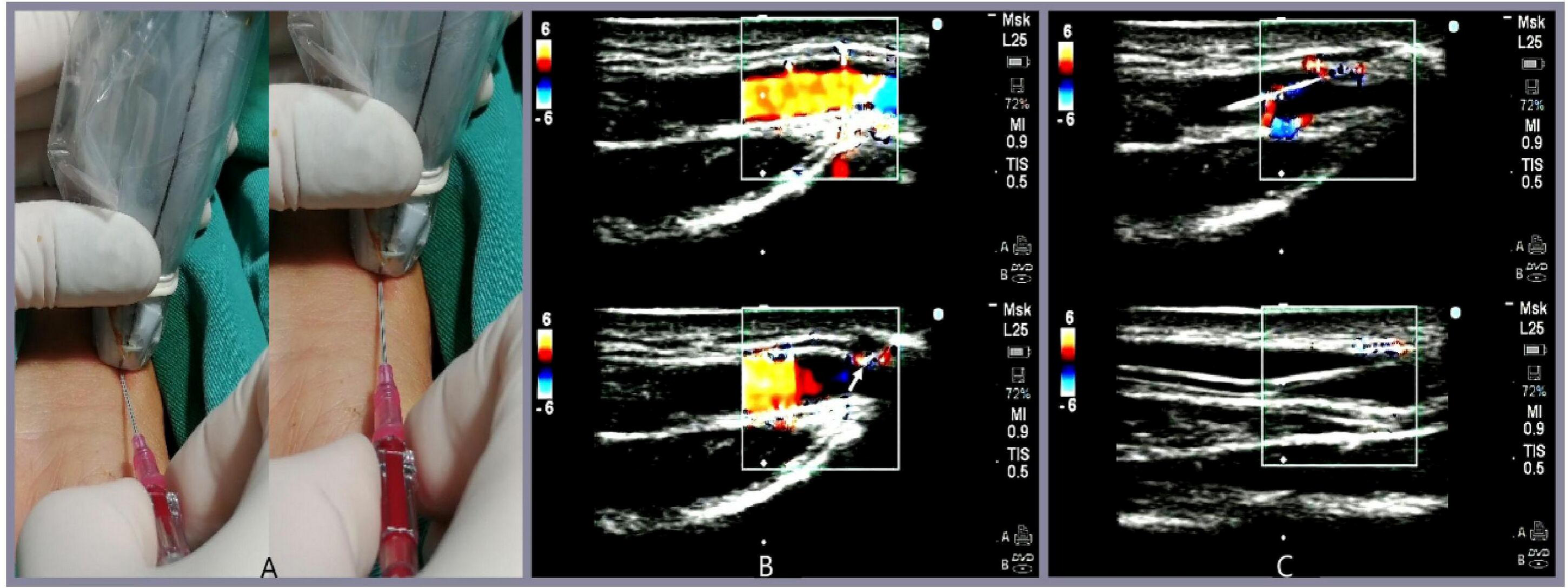
The modified long-axis in-plane ultrasound technique (M-LAINUTP) approach. A. The black line on the probe is the centre line to guide radial artery cannulation. B. The needle (the white arrow) had been inserted into the anterior wall of the radial artery, the vascular wall was compressed, and the blood flow signal was weakened on the ultrasound image. C. The image of the needle or cannula in the radial artery was clearly displayed on the ultrasound

In palpation, the operator palpated the radial artery pulse with the left index and middle fingers. The strongest pulse point was identified as the puncture point. The needle and cannula were advanced toward the radial artery at an angle of approximately 15°–30°. Once blood appeared in the hub, the angle of needle decreased slightly, and the needle was advanced approximately 3mm. If blood continued to flow into the hub, the cannula was pushed into the artery.

The data were collected by other anesthesiologists who were not associated with the performance of the procedure, including age, height, body mass index, sex, and history of peripheral vascular disease (PVD). Twenty-four hours and three days after surgery were the follow-up times, during which data were recorded by anesthesiologists who were not aware of the grouping. The primary investigators concurrently assessed the same endpoints for a sample of 10% of study cannulations to confirm the accuracy of the data. Measurement of Outcomes

The primary outcome was the success of the first cannulation which was defined as the successful insertion of the cannula with one skin puncture for the first time and the acquisition of arterial waveforms. Cannula insertion failure was defined as greater than 3 cannulation attempts for a single arm or an attempted cannulation time more than 5 minutes.^4^.

Secondary outcomes included the cannulation time (seconds), the number of attempts, complications, etc. The cannulation time was defined as the interval between the first skin penetration and confirmation of the arterial waveform on the monitor. The number of attempts was defined as the number of skin perforations caused by the puncture needle.

### 2.1. Statistical Analysis

According to a previous study,^9^ the success rate of puncture was 68% in the palpation group and 92% in the ultrasound group. With an expected 10% loss rate per group, we calculated a sample size of 288 patients by the Z-pooled normal approximation method, and 96 patients would receive radial artery cannulation from each operator. All the data were processed by SPSS software (Version 24.0, IBM, USA). The measured data are expressed as the mean±standard deviation. First, normality and variance homogeneity were tested, and the t test was used for comparisons between groups. If the variance was uneven, the two-sample rank-sum test was used to compare groups. The count data were tested by the χ^2^ test. The 95% confidence intervals were calculated by the Woolf method. A two-tailed p <0.05 was considered statistically significant.

## 3. Results

In this study, three patients were excluded, so 285 patients were randomized into the final analysis. Figure 2 illustrates participant recruitment and group allocation. Reasons for exclusion included the reluctance to randomize (M-LAINUT, n=1), the attending anesthesiologist decided not to insert the cannula into the radial artery (C-PT, n=1), and the loss of medical records (C-PT, n=1). General basic characteristics were similar between the two groups (Table 1).

**Table 1.**
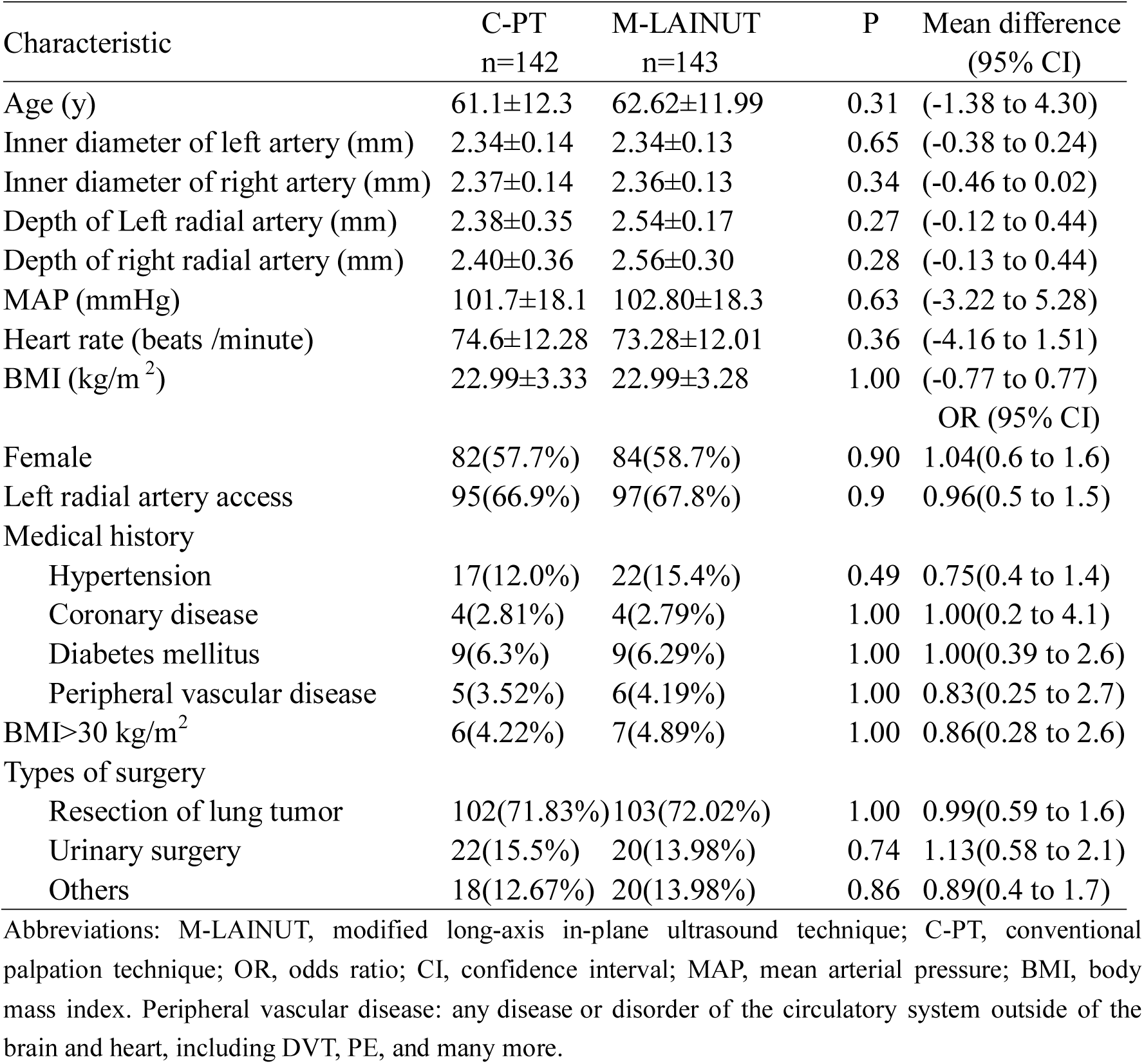
Baseline Patient Characteristics

**Figure 2.**
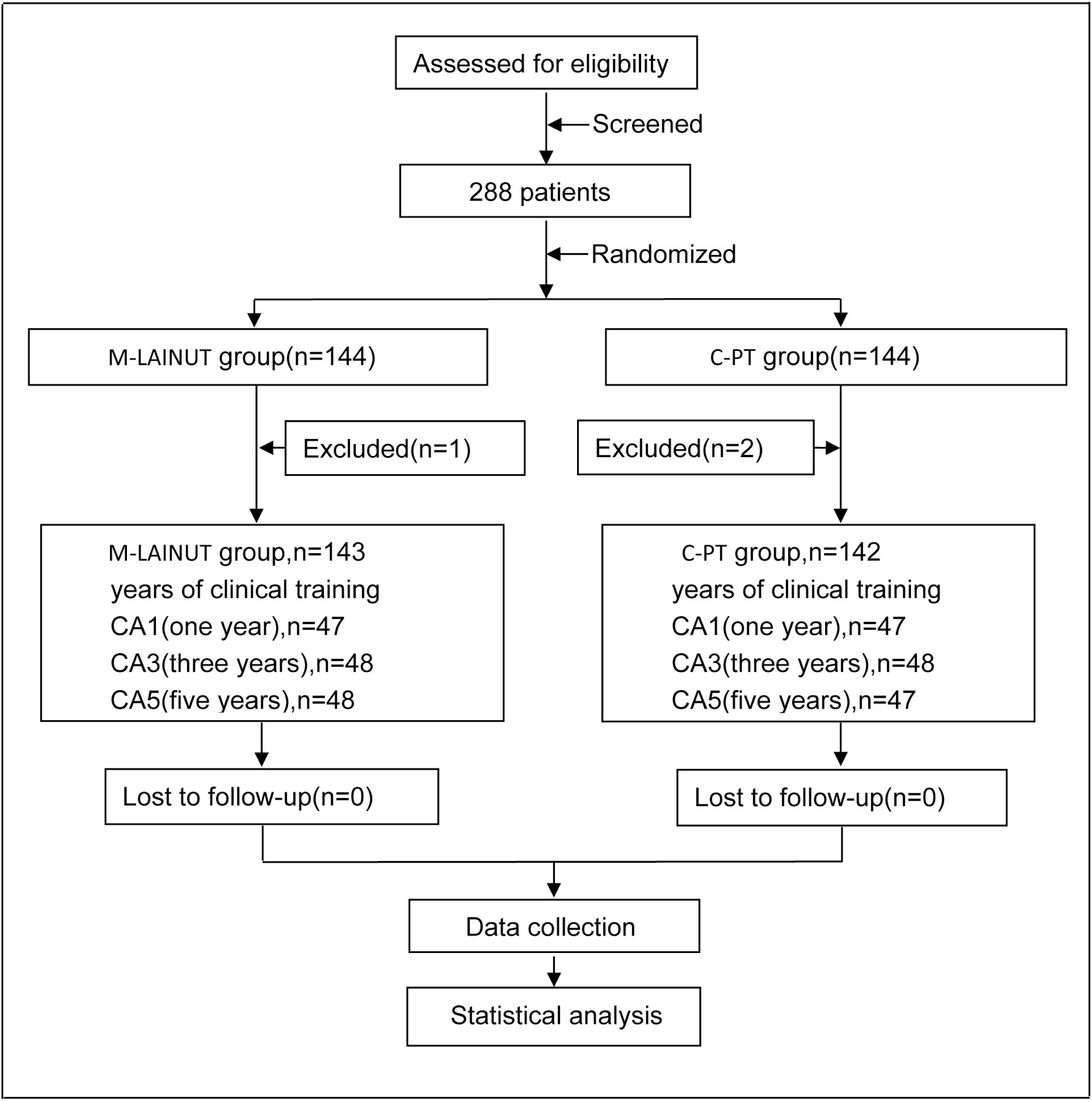
Flow diagram of patient recruitment and randomization.

Cannulation success rates on the first attempt and total attempts in the M-LAINUT group were 91.6% and 97.9%, respectively, which were significantly higher than the 57.7% and 84.5% in the C-PT group (p <0.001, Table 2). The cannula insertion failure rate in the C-PT group was 15.5%, which was significantly higher than the 2.1% in the M-LAINUT group (p <0.001, Table 2).

**Table:2.** Comparison of the success rate of puncture between the two groups

| Number attempts<br>(<5 min) | C-PT<br>(N=142)<br>n (%) | M-LAINUT<br>(N=143)<br>n (%) | P | OR | 95% CI |
| --- | --- | --- | --- | --- | --- |
| 1<sup>a</sup> | 82 (57.7) | 131 (91.6) | <0.001 | 7.9 | 4.0 to 15.7 |
| 2 | 18 (12.7) | 8 (4.4) |  |  |  |
| 3 | 20 (14.1) | 1 (0.7) |  |  |  |
| Total success rate<sup>a</sup> | 120 (84.5) | 140 (97.9) | <0.001 | 8.5 | 2.5 to 29.2 |
| Rate of insertion failure<sup>b</sup> | 22 (15.5) | 3 (2.1) | <0.001 | 0.1 | 0.03 to 0.4 |

Abbreviations: M-LAINUT, modified long-axis in-plane ultrasound technique; C-PT, conventional palpation technique; OR, odds ratio; CI, confidence interval.
<sup>a</sup> P values and 95% CIs are calculated from the $\chi^2$ test.
<sup>b</sup> P values and 95% CIs are calculated from Fisher's exact test.

The total cannulation time of the C-PT group was longer than that of the M-LAINUT group (Z=-4.08, p <0.001, figure 3). The data of total cannulation time were nonnormally distributed and the variance was uneven, the two-sample rank-sum test was used for statistical processing. Our results showed a significant reduction in the number of attempts required to cannulate the radial artery with M-LAINUT versus C-PT (mean: 1.11±0.39 vs 1.72±0.89, p <0.001, figure 4).

**Figure 3.**
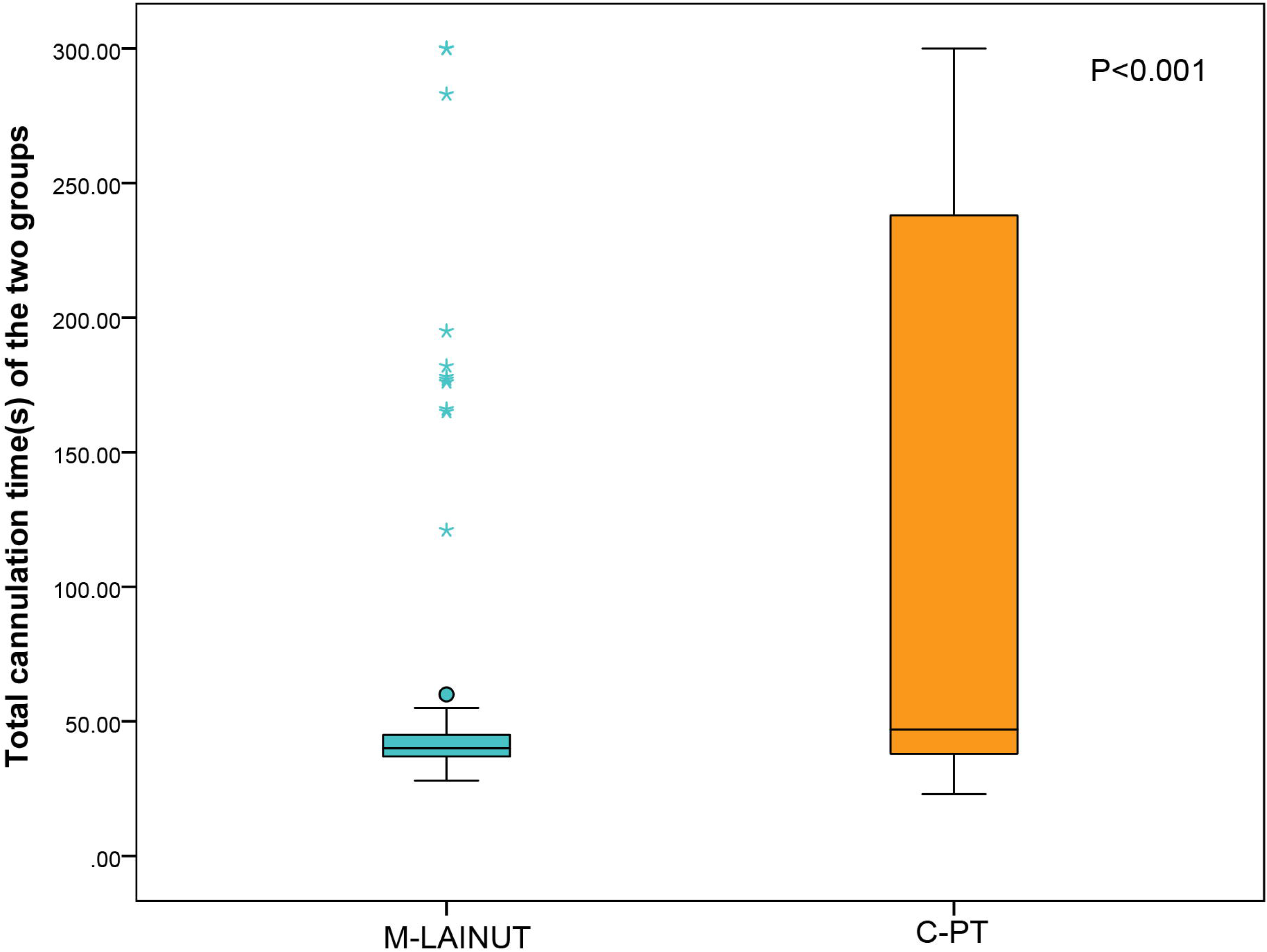
Total cannulation time in the two groups.

**Figure 4.**
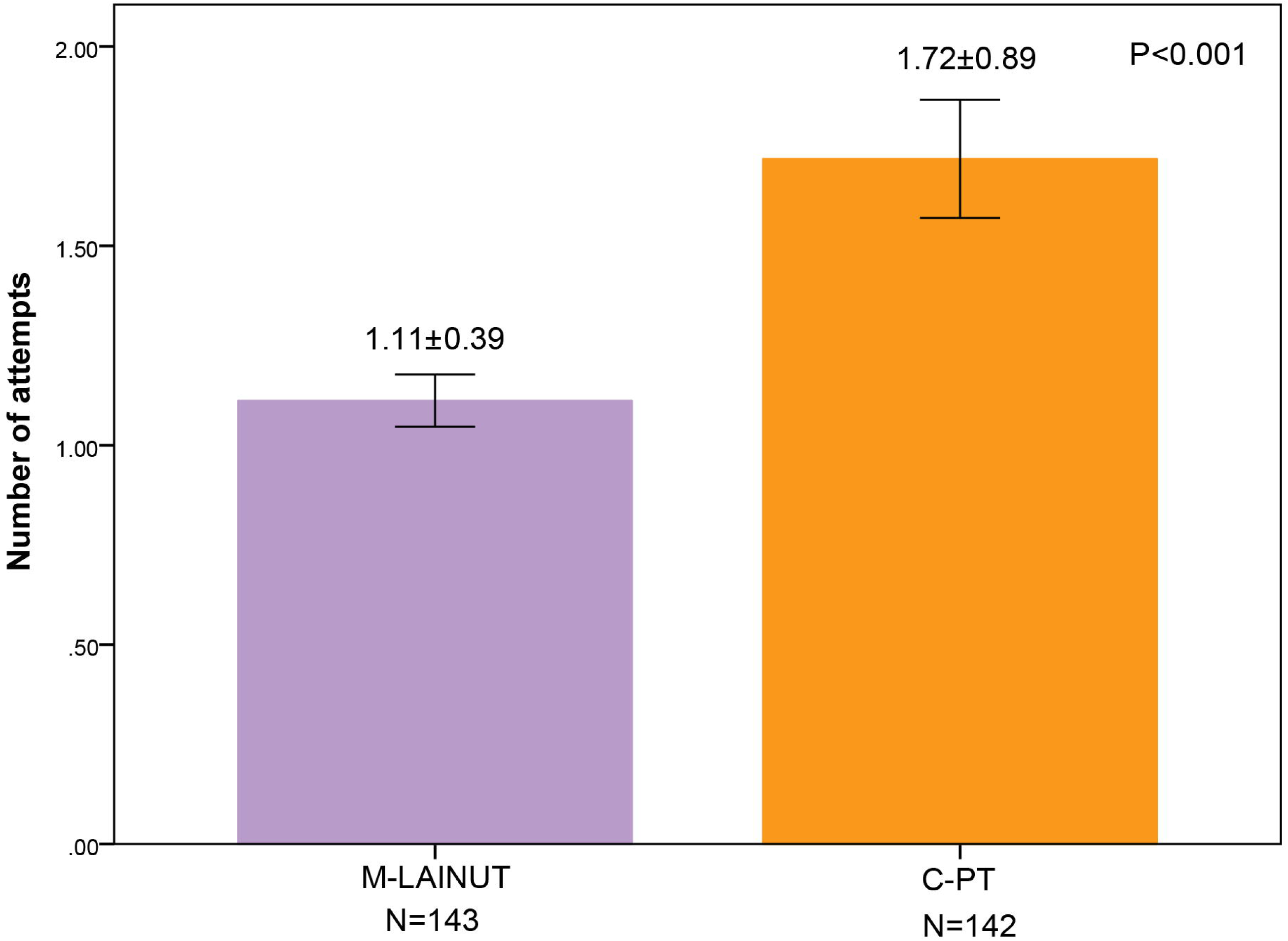
Number of attempts.

The incidence of hematoma in the C-PT group was 19.7%, which was significantly higher than the 2.8% in the M-LAINUT group (p <0.001, OR, 8.54, 95% CI,2.9 to 25.0, table 4). We found that hematoma caused by puncture was a high risk factor for cannulation failure in both groups (p <0.001; OR, 118; 95% confidence interval, 34–405, figure 5).

**Table:3.** Comparison of the rate of success on the second and third attempts combined C-PT M-LAINUT

| Subgroup | C-PT | M-LAINUT | P | OR | 95% CI |
| --- | --- | --- | --- | --- | --- |
|  | n/N (%) | n/N(%) |  |  |  |
| CA1 | 15/47 (31.9)<sup>a</sup> | 5/47 (10.63) | <0.001 | 0.06 | 0.02 to 0.17 |
| CA3 | 16/48 (33.3)<sup>b</sup> | 3/48 (6.25) | <0.001 | 0.03 | 0.01 to 0.12 |
| CA5 | 7/47 (14.9) | 1/48 (2.1) | <0.001 | 0.004 | 0.0001 to 0.03 |
| Total | 38/142 (26.76) | 9/143 (6.3) | <0.001 | 0.03 | 0.01 to 0.05 |

Abbreviations: CA, clinical anesthetists, who are divided into three categories according to the year of clinical training in anesthesia after graduation from medical college: one year (CA1), three years (CA3), five years (CA5).
P values and ORs (95% CIs) are calculated from Fisher's exact test.

**Table:4.**
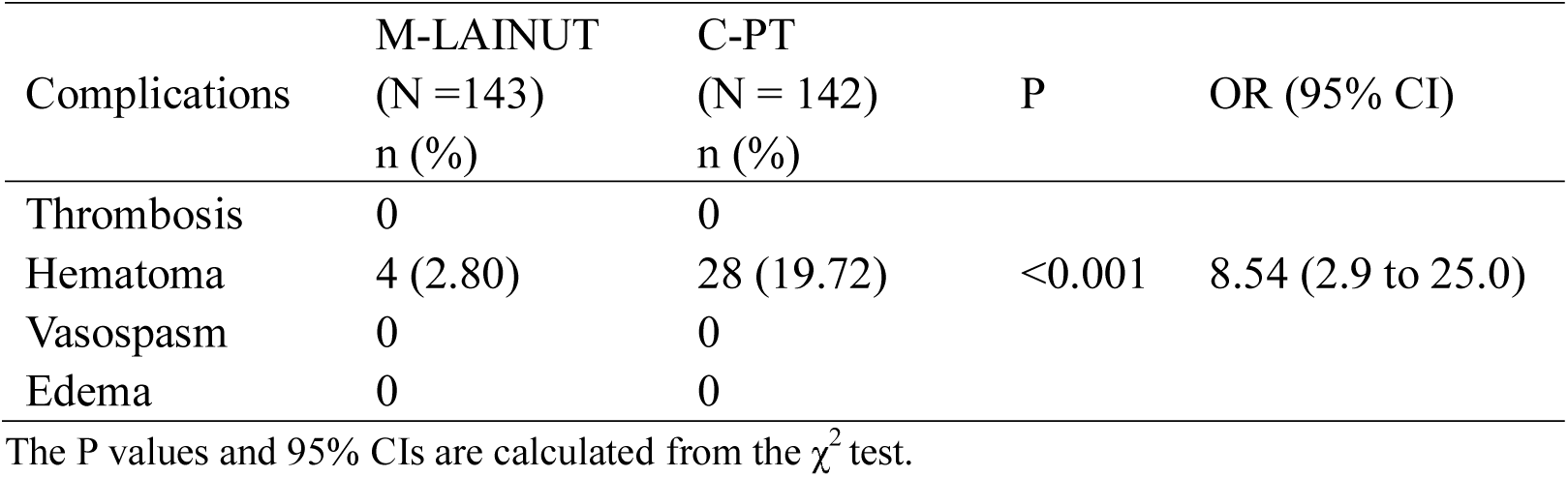
Complications Recorded During Arterial Cannulation

| Complications | M-LAINUT<br>(N =143)<br>n (%) | C-PT<br>(N = 142)<br>n (%) | P | OR (95% CI) |
| --- | --- | --- | --- | --- |
| Thrombosis | 0 | 0 |  |  |
| Hematoma | 4 (2.80) | 28 (19.72) | <0.001 | 8.54 (2.9 to 25.0) |
| Vasospasm | 0 | 0 |  |  |
| Edema | 0 | 0 |  |  |

The P values and 95% CIs are calculated from the $\chi^2$ test.

**Figure 5.**
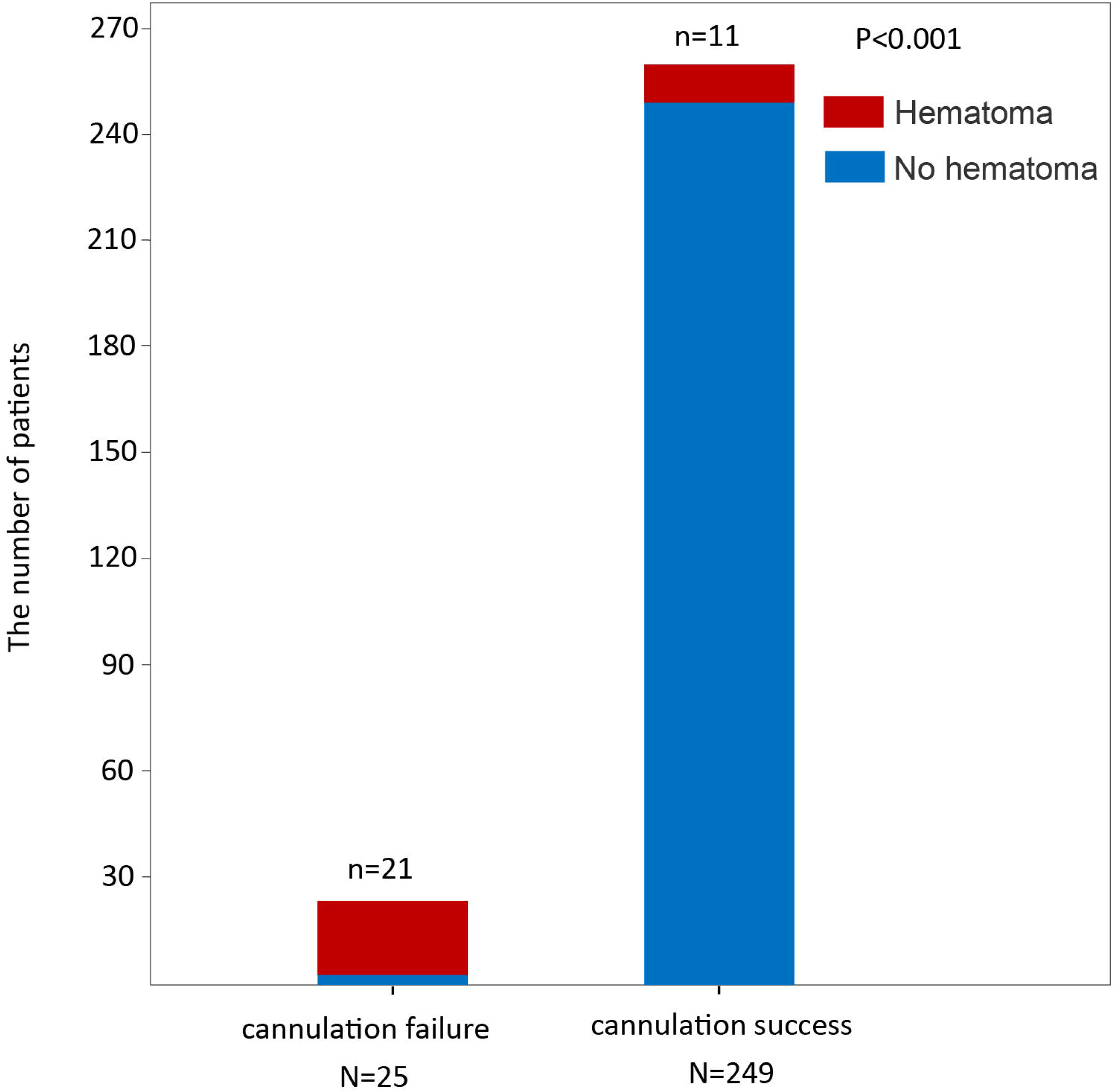
Relationship between hematoma and cannulation failure in the two groups

### Subgroup Analyses

In three subgroups (CA1, CA3, CA5), the success rate of the first attempt in the M-LAINUT group was significantly higher than that in the C-PT group (p<0.001, figure 6).

**Figure 6.**
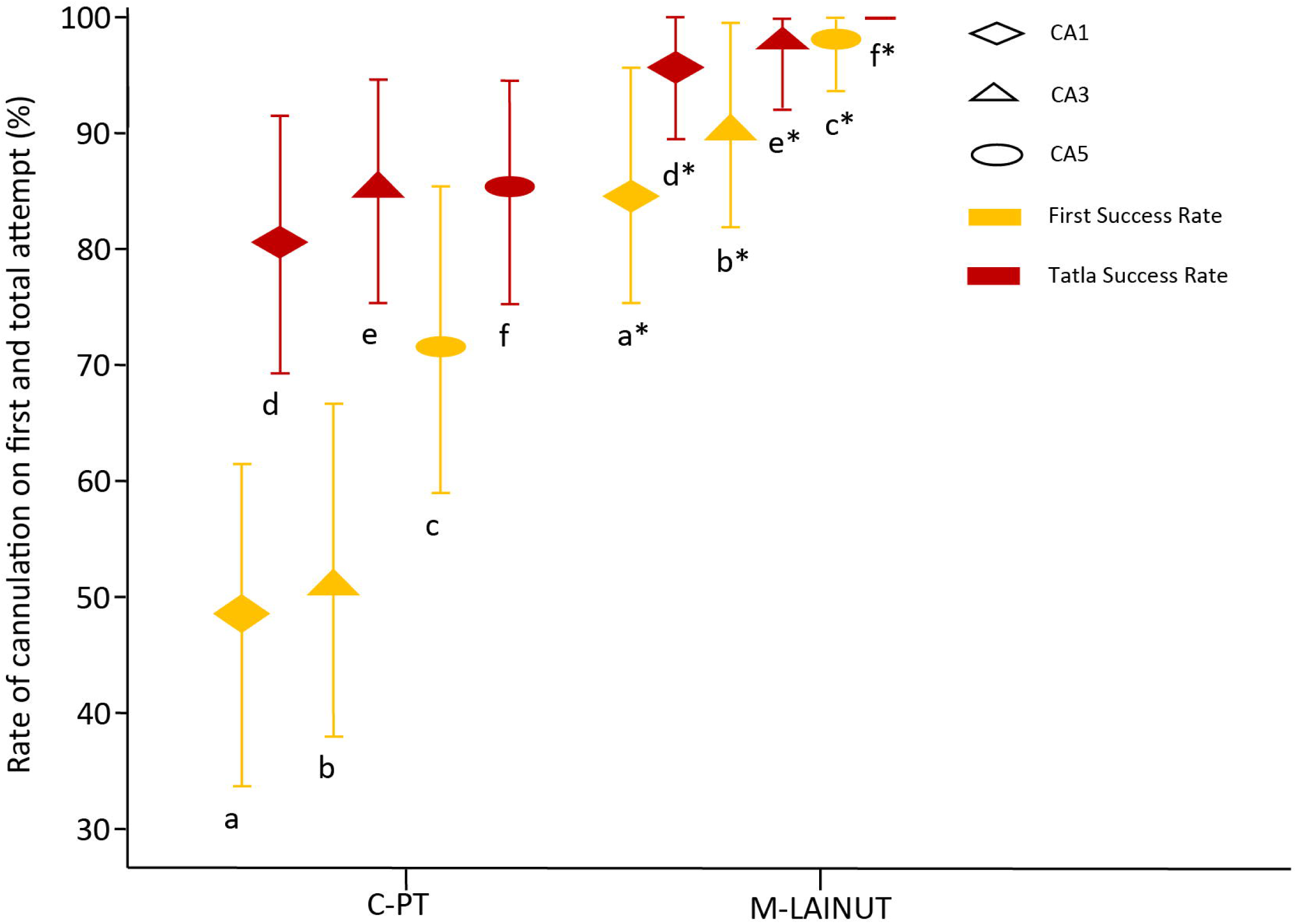
First-attempt success rate and total success rate in subgroups.

The overall success rate of M-LAINUT in the CA1 and CA3 groups was higher than that in C-PT group, but not significantly (p> 0.05, figure 6). In the CA5 group, the overall success rate of M-LAINUT was significantly higher than that of the C-PT group (p <0.05, figure 6).

The first and total success rates of C-PT in the CA5 group were 72.3% and 87.2%, respectively, which were lower than those of M-LAINUT in the CA1 group (85.1% and 95.7%). However, there was no statistically significant difference between the two groups (p>0.05, figure 6).

In the M-LAINUT group, the first-attempt success rate of CA5 was 97.9%, which was significantly higher than the 85.1% of the CA1 group (p<0.05, figure 6). However, there was no statistically significant difference in the total cannulation success rate between the two groups (p>0.05, figure 6).

In each subgroup, the second and third attempt success rate of cannulation in the C-PT group was significantly higher than that in the M-LAINUT group (p <0.001, Table 3). We also found that the C-PT success rate on the second and third attempts in the CA5 group was significantly lower than that of CA1 or CA3 (p ^a^ < 0.001, p ^b^ < 0.001, Table 3).

## 4. Discussion

In this study, we found that the use of the M-LAINUT significantly increased the first and total cannulation success rates, reduced the number of attempts and shortened the total successful cannulation time compared with the traditional palpation technique. Moreover, we also found that for the young anesthesiologists who lacked ultrasound experience, M-LAINUT significantly improved the total success rate. For an experienced anesthesiologist, the use of M-LAINUT can improve the first-attempt success rate. In addition, we found that hematoma caused by puncture was a high risk factor for the failure of cannulation.

Obtaining a long axis view may be challenging since the thickness of the ultrasonic beam is approximately 1-2 mm. However, after obtaining a short axis view of the artery, the US probe was rotated 90°, keeping the arterial image in the centre of the ultrasound screen, which can help to identify and obtain the long axis view of the radial artery.^10^ Color mode can effectively capture the radial artery blood flow signal, and by analyzing the intensity of the blood flow signal and the diameter of artery, the clinician can adjust the ultrasound plane to perfectly align it with the long axis of the blood vessel.

In our study, the centre line of the ultrasonic probe is designed to guide the needle to align with the ultrasonic plane, which can increase the visibility of needle and greatly improve the accuracy of puncture. The use of the special puncture posture and needle-holding mode could facilitate alignment of the needle with the centre line of probe. The wrist joint angle of the patient was adjusted to 45°, which maximizes the radial artery height and decreases the distance between the radial artery and skin significantly.^11^ The results also confirm that the M-LAINUT improved the success rates of the first and total cannulation, reduced the failure rate.

In our study, second and third puncture attempts may result in hematoma (the incidence of hematoma was higher in the C-PT group) and increase the difficulty of subsequent cannulation, which may lead to the longer total cannulation time in the C-PT group.

For CA1 and CA3, the second and third attempt cannulation success rate of the C-PT group was higher than that of the M-LAINUT group. To some extent, the second to third attempt success rate of C-PT compensated for the deficiency in the first cannulation success rate, which may have caused the small difference in the total cannulation success rate between the two groups. However, in the C-PT group, the success rate of the second and third cannulations in the CA5 group was significantly lower than that of CA1 and CA3, which may be the reason why the total success rate of M-LAINUT in the CA5 group was higher than that of C-PT group.

Regarding the learning curve of ultrasound-guided and operator experience playing an important role in successful cannulation,^12,13^ we limited the participants to three anesthesiologists with varying levels of experience. Despite the first-pass rate of ultrasound-guided radial artery cannulation was higher than that of palpation approach,^14,15^ it was still a low rate, ranging from 51%--76%^4,10,13,16,17^ and multiple attempts may cause serious complications, including vasospasm, hematoma.^18,19^ The M-LAINUT can increase the first attempt success rate to more than 90%.

It has been reported that compared with the palpation technique, ultrasound guidance improved the first and total success rates of radial artery cannulation, shortened the cannulation time^3,20^ and reduced the incidence of complications^21^ which is in concordance with the results of our study. However, a meta-analysis^15,22^ found that ultrasound guidance did not increase the total success rate of radial artery cannulation, which is inconsistent with our findings, the reason for which may be related to the different ultrasound technology, patients population, operators experience.

A study^10^ using modified short-axis out-of-plane ultrasound to guide radial artery cannulation showed that the success rate of the first cannulation was 88.9%, which was similar to that in our study (91.6%), but the total success rate (100%) was higher than that in our study (97.9%). The reason could be that, in addition to the different ultrasound guidance techniques used, the operators were different: an experienced anesthesiologist had used the ultrasound-guided technique for approximately 200 procedures. Our study involved three anesthesiologists with different clinical experience who underwent short-term training of ultrasound-guided artery cannulation.

The technique of ultrasound-guided dynamic needle tip positioning has significantly improved the success rate of radial artery catheterization.^5,6,21^ Another study^18^ using ultrasound-guided dynamic tip localization for radial artery cannulation showed that the average diameter of the radial artery was 2.8 mm, with a first success rate of 83% and a total success rate of 89.3%, which was inconsistent with our study. This may be related to the number of operators and the different training and experience of the operators; 41 residents and faculty members performed radial artery cannulation, which may have reduced the success rate.

Research shows^4^ that inexperience may have prevented trainees from realizing the full benefit of ultrasound, the effect of which was greatest for the most experienced trainees, which is consistent with our study.

Quan Z F et al.^10^ observed an incidence of hematoma in the ultrasound group of 14.8%, which is higher than the M-LAINUT group in our study. One possible reason is that the long-axis plane ultrasound in this study may have improved the visibility of the puncture tip,^16^ which reduced the damage to the radial artery caused by the needle.^23^

After familiarity with the use of the US-guided approach, we presume that the M-LAINUT can be easily extrapolated to central venous cannulation, nerve block anesthesia in all the area of clinical settings, including intensive care units and operating rooms. ^24^

### Study limitations

It may be more suitable for cases of difficult cannulation of the radial artery, considering that the ultrasound technique may increase the cost to the patient. For some patients with high risk factors for difficult cannulation of radial artery, such as artery diameter less than 2.2 mm, critically ill children,^25^ infants,^26^ it is not clear whether this technique has advantages over traditional ultrasound and palpation techniques. Three anesthesiologists in the CA1, CA3 and CA5 groups were involved in the study, so the Hawthorne effect could not be completely excluded. To further evaluate the advantage of the M-LAINUT, future study is needed regarding difficult case, multicenter randomized controlled trial and comparing with short axis ultrasonography.

In conclusion, the modified long-axis in-plane ultrasound technique, when performed by operators with different experiences, demonstrates some clinical advantages for radial artery cannulation in adults, with a higher success rate of cannulation, fewer attempts and less time.

## Data Availability

The data used to support the findings of this study are included within the article.

## Abbreviations

ICU: intensive care unit
M-LAINUT: modified long-axis in-plane ultrasound technique
C-PT: the conventional palpation technique
CFM: color doppler mode
OR: odds ratio
CI: confidence interval
MAP: mean arterial pressure
BMI: body mass index
PVD: peripheral vascular disease
CA: clinical anesthetists
SD: standardized difference

## Acknowledgements

The authors would like to thank the anesthesiologists, patients and nurses of the anesthesia operating department.

